# Investigating Chemokine-Induced NF-κB Activation in Retinal Cells and Its Contribution to Age-Related Macular Degeneration Pathogenesis: A Systematic Review

**DOI:** 10.1101/2024.09.07.24313230

**Authors:** Ifrah Siddiqui, Nabeel Ahmad Khan, Fatima Ahmad, Maham Nawaz, Almas Naeem, Muhammad Usaid Khalid, Usman Haroon Mirza, Hamza Ali Danish, Syed Saqib Khalid

## Abstract

**Objective:** To investigate the roles of chemokines in activating the NF-κB signaling pathway in retinal pigment epithelium (RPE) and photoreceptor cells, and their contribution to the pathogenesis of age-related macular degeneration (AMD).

**Background:** AMD is a leading cause of vision loss in older adults, driven by chronic inflammation and oxidative stress. The NF-κB transcription factor plays a key role in regulating these processes, with various chemokines, such as CCL2 and CX3CL1, influencing NF-κB activation. Despite advances in treatment, a deeper understanding of how chemokines affect NF-κB activation in RPE and photoreceptor cells remains critical for developing effective therapies. This study seeks to address this gap and improve AMD management.

**Methods:** A comprehensive search of databases, including PubMed, MEDLINE, and Google Scholar, was conducted to identify relevant studies on the roles of chemokines in activating NF-κB signaling in RPE and photoreceptor cells. The search covered chemokines such as CCL2 (MCP-1), CX3CL1 (Fractalkine), CCL3 (MIP-1α), CCL5 (RANTES), CXCL8 (IL-8), CXCL10 (IP-10), CXCL1 (GRO-α), CXCL12 (SDF-1), CCL11 (Eotaxin), CXCL16, CXCL9 (MIG), and CXCL11 (I-TAC). Studies were systematically reviewed following PRISMA guidelines to assess the involvement of these chemokines in NF-κB activation.

**Results:** The analysis revealed that chemokines, including CCL2, CX3CL1, CCL3, and CCL5, significantly activated NF-κB in RPE and photoreceptor cells, leading to increased inflammation and apoptosis through enhanced cytokine production and reactive oxygen species (ROS) generation. Additionally, CXCL8, CXCL10, CXCL1, and CXCL12 triggered NF-κB activation, contributing to oxidative stress and extracellular matrix (ECM) remodeling, which disrupts retinal structure and function. Other chemokines, such as CCL11, CXCL16, CXCL9, and CXCL11, sustained chronic inflammation and modulated matrix metalloproteinases (MMPs), further implicating them in AMD progression.

**Conclusion:** Chemokines, including CCL2, CX3CL1, CCL3, CCL5, CXCL8, CXCL10, CXCL1, CXCL12, CCL11, CXCL16, CXCL9, and CXCL11, activate the NF-κB pathway in RPE and photoreceptor cells, driving chronic inflammation, oxidative stress, and ECM remodeling in AMD. These findings highlight potential therapeutic targets to mitigate disease progression.

## Background

Age-related macular degeneration (AMD) is a leading cause of vision loss among the elderly population worldwide, characterized by progressive degeneration of the retinal pigment epithelium (RPE) and photoreceptor cells. AMD manifests in two forms: dry (atrophic) and wet (neovascular) [<u>1</u>]. Despite significant advancements in treatment, including anti-VEGF therapies for wet AMD, there remains an urgent need to understand the underlying mechanisms driving AMD to develop more effective interventions for both forms of the disease [<u>6</u>].

Recent studies have highlighted the role of inflammatory processes in AMD pathogenesis, with a particular focus on the activation of the nuclear factor kappa B (NF-κB) signaling pathway. NF-κB, a key transcription factor, is involved in regulating inflammatory responses, cellular survival, and stress responses. Dysregulation of NF-κB signaling has been implicated in the chronic inflammation and oxidative stress observed in AMD [<u>2</u>].

Chemokines are key modulators of inflammation and immune cell recruitment, influencing the NF-κB pathway in various tissues. In the context of AMD, several chemokines—such as CCL2 (MCP-1), CX3CL1 (Fractalkine), CCL3 (MIP-1α), CCL5 (RANTES), CXCL8 (IL-8), CXCL10 (IP-10), CXCL1 (GRO-α), CXCL12 (SDF-1), CCL11 (Eotaxin), CXCL16, CXCL9 (MIG), and CXCL11 (I-TAC)—have been implicated in driving retinal inflammation and damage [<u>3</u>]. However, the specific roles of these chemokines in NF-κB activation and their contributions to AMD remain inadequately explored [<u>7</u>].

Understanding how these chemokines influence NF-κB signaling in RPE and photoreceptor cells is crucial for elucidating the mechanisms of AMD pathogenesis [<u>4</u>]. This study aims to address these gaps by investigating the molecular mechanisms through which chemokine-induced NF-κB activation affects cellular processes and contributes to AMD. Insights gained from this research could inform the development of novel therapeutic strategies targeting inflammation and oxidative stress, potentially leading to better management and prevention of AMD [<u>5</u>].

## Methods

### Aim of the Study

This study aims to investigate the role of specific chemokines—CCL2 (MCP-1), CX3CL1 (Fractalkine), CCL3 (MIP-1α), CCL5 (RANTES), CXCL8 (IL-8), CXCL10 (IP-10), CXCL1 (GRO-α), CXCL12 (SDF-1), CCL11 (Eotaxin), CXCL16, CXCL9 (MIG), and CXCL11 (I-TAC)—in the activation of the NF-κB signaling pathway in retinal pigment epithelium (RPE) and photoreceptor cells. The study focuses on elucidating the molecular mechanisms involved and how this activation influences cellular processes, including inflammation, oxidative stress, apoptosis, and extracellular matrix remodeling, contributing to the pathogenesis of Age-related Macular Degeneration (AMD).

### Research Question

How do the chemokines CCL2, CX3CL1, CCL3, CCL5, CXCL8, CXCL10, CXCL1, CXCL12, CCL11, CXCL16, CXCL9, and CXCL11 activate the NF-κB signaling pathway in retinal pigment epithelium (RPE) and photoreceptor cells, and what are the molecular mechanisms through which this activation influences cellular processes that contribute to the development of Age-related Macular Degeneration (AMD)?

### Search Focus

A comprehensive literature search was conducted using multiple databases, including PUBMED, MEDLINE, and Google Scholar, along with open-access and subscription-based journals. There were no date restrictions for published articles. The search focused on the following chemokines and their involvement in NF-κB signaling in RPE and photoreceptor cells:

- CCL2 (MCP-1)
- CX3CL1 (Fractalkine)
- CCL3 (MIP-1α)
- CCL5 (RANTES)
- CXCL8 (IL-8)
- CXCL10 (IP-10)
- CXCL1 (GRO-α)
- CXCL12 (SDF-1)
- CCL11 (Eotaxin)
- CXCL16
- CXCL9 (MIG)
- CXCL11 (I-TAC)

The literature was screened and selected based on relevance to the study’s objectives. Literature search began in October 2021 and ended in March 2024. An in-depth investigation was conducted during this duration based on the parameters of the study as defined above. During revision, further literature was searched and referenced until September 2024. The literature search and all sections of the manuscript were checked multiple times during the months of revision (April 2024 – September 2024) to maintain the highest accuracy possible. This approach allowed for a comprehensive investigation of the role of these chemokines in NF-κB pathway activation and their contribution to AMD pathogenesis, adhering to the PRISMA guidelines for systematic reviews.

### Search Queries/Keywords

1. **General Terms:**

"Age-related Macular Degeneration" OR "AMD" "Retinal Pigment Epithelium" OR "RPE" "Photoreceptor Cells"
"NF-κB signaling pathway"
2. **Chemokines:**

"CCL2" AND "AMD"
"CX3CL1" AND "NF-κB signaling"
"CCL3" AND "RPE"
"CCL5" AND "photoreceptor cells" "CXCL8" AND "oxidative stress" "CXCL10" AND "apoptosis"
"CXCL1" AND "extracellular matrix remodeling" "CXCL12" AND "inflammation"
"CCL11" AND "AMD"
"CXCL16" AND "NF-κB signaling"
"CXCL9" AND "RPE"
"CXCL11" AND "NF-κB activation"

Boolean operators (AND, OR) were utilized to construct search queries that combined these chemokines with terms related to AMD, NF-κB signaling, and key cellular processes. This search strategy was designed to capture relevant studies focusing on the molecular mechanisms involved in chemokine-driven NF-κB activation and its contribution to AMD.

### Objectives of the Searches

- To determine the role of specific chemokines in activating the NF-κB signaling pathway in RPE and photoreceptor cells.
- To elucidate the molecular mechanisms underlying chemokine-driven NF-κB activation.
- To investigate how this activation influences inflammation, oxidative stress, apoptosis, and extracellular matrix remodeling.
- To assess the contribution of these processes to the pathogenesis of Age-related Macular Degeneration (AMD).

### Screening and Eligibility Criteria

#### Initial Screening

Articles were initially screened based on titles and abstracts to identify those directly addressing the role of chemokines in NF-κB signaling, particularly in the context of RPE and photoreceptor cells.

#### Full-Text Review

Following initial screening, full-text articles were reviewed to ensure they met the study’s criteria for relevance and quality. Only studies providing detailed insights into the molecular mechanisms of chemokine-driven NF-κB activation and its impact on cellular processes related to AMD were included.

#### Data Extraction

Data extraction focused on key findings related to the role of CCL2, CX3CL1, CCL3, CCL5, CXCL8, CXCL10, CXCL1, CXCL12, CCL11, CXCL16, CXCL9, and CXCL11 in NF-κB activation, as well as their influence on inflammation, oxidative stress, apoptosis, and extracellular matrix remodeling in AMD.

### Inclusion and Exclusion Criteria

#### Inclusion Criteria

- Studies focusing on the role of CCL2, CX3CL1, CCL3, CCL5, CXCL8, CXCL10, CXCL1, CXCL12, CCL11, CXCL16, CXCL9, and CXCL11 in activating the NF-κB signaling pathway in RPE and photoreceptor cells.
- Research that explores the molecular mechanisms linking NF-κB activation to inflammation, oxidative stress, apoptosis, and extracellular matrix remodeling.
- Articles that investigate the contribution of these processes to AMD pathogenesis.

#### Exclusion Criteria

- Studies that do not directly focus on the NF-κB signaling pathway or the specified chemokines.
- Research that lacks detailed mechanistic insights into chemokine-driven NF-κB activation.
- Articles that focus on unrelated diseases or cellular processes.
- Non-English language publications and conference abstracts or unpublished studies.

### Rationale for Screening and Inclusion

The chemokines under investigation were selected based on their known roles in inflammation and immune regulation, particularly in ocular tissues. Each chemokine was reviewed for its potential to activate the NF-κB signaling pathway and influence cellular processes relevant to AMD:

- **CCL2 (MCP-1)**: Known for recruiting monocytes and macrophages, contributing to chronic inflammation in AMD.
- **CX3CL1 (Fractalkine)**: Modulates microglial activity, influencing neuroinflammation and NF-κB activation.
- **CCL3 (MIP-1α)**: Promotes inflammatory responses and may impact RPE and photoreceptor cell function.
- **CCL5 (RANTES)**: Enhances leukocyte recruitment, potentially contributing to retinal inflammation.
- **CXCL8 (IL-8)**: Induces oxidative stress and inflammation, key processes in AMD progression.
- **CXCL10 (IP-10)**: Involved in apoptotic pathways and immune responses in retinal cells.
- **CXCL1 (GRO-α)**: Influences extracellular matrix remodeling, affecting retinal structural integrity.
- **CXCL12 (SDF-1)**: Plays a role in tissue repair and inflammation, impacting AMD development.
- **CCL11 (Eotaxin)**: Linked to chronic inflammation, which may exacerbate retinal degeneration.
- **CXCL16**: Modulates immune responses and interacts with NF-κB signaling in retinal tissues.
- **CXCL9 (MIG)** and **CXCL11 (I-TAC)**: Both are involved in pro-inflammatory signaling pathways, potentially contributing to AMD-related pathology.

### Assessment of Article Quality and Potential Biases

Ensuring the quality and minimizing potential biases of the selected articles were crucial to ensuring the reliability of the study’s conclusions.

**Quality Assessment**: Articles were evaluated for their methodological rigor, including experimental design, data analysis, and reproducibility. Peer-reviewed articles were prioritized, ensuring that the included studies adhered to high standards of scientific inquiry.

**Bias Assessment**: Several potential biases were considered:

- **Publication Bias**: A comprehensive search strategy, including negative and positive results, was adopted to minimize publication bias.
- **Selection Bias**: Predefined inclusion criteria were strictly applied to ensure objectivity in article selection.
- **Reporting Bias**: Articles were checked for completeness and consistency, and multiple reviews were conducted to minimize reporting bias.

### Language and Publication Restrictions

Only English-language publications were included. No restrictions were placed on the date of publication to ensure a broad range of studies were considered. Unpublished studies, including conference abstracts, were excluded to prioritize peer-reviewed and validated research findings.

## Results

A total of 2205 articles were identified using database searching, and 2101 were recorded after duplicates removal. 1789 were excluded after screening of title/abstract, 176 were finally excluded, and 5 articles were excluded during data extraction. These exclusions were primarily due to factors such as non-conformity with the study focus, insufficient methodological rigor, or data that did not align with our research questions. Finally, 131 articles were included as references. PRISMA Flow Diagram is Fig 1.

**Fig 1.**
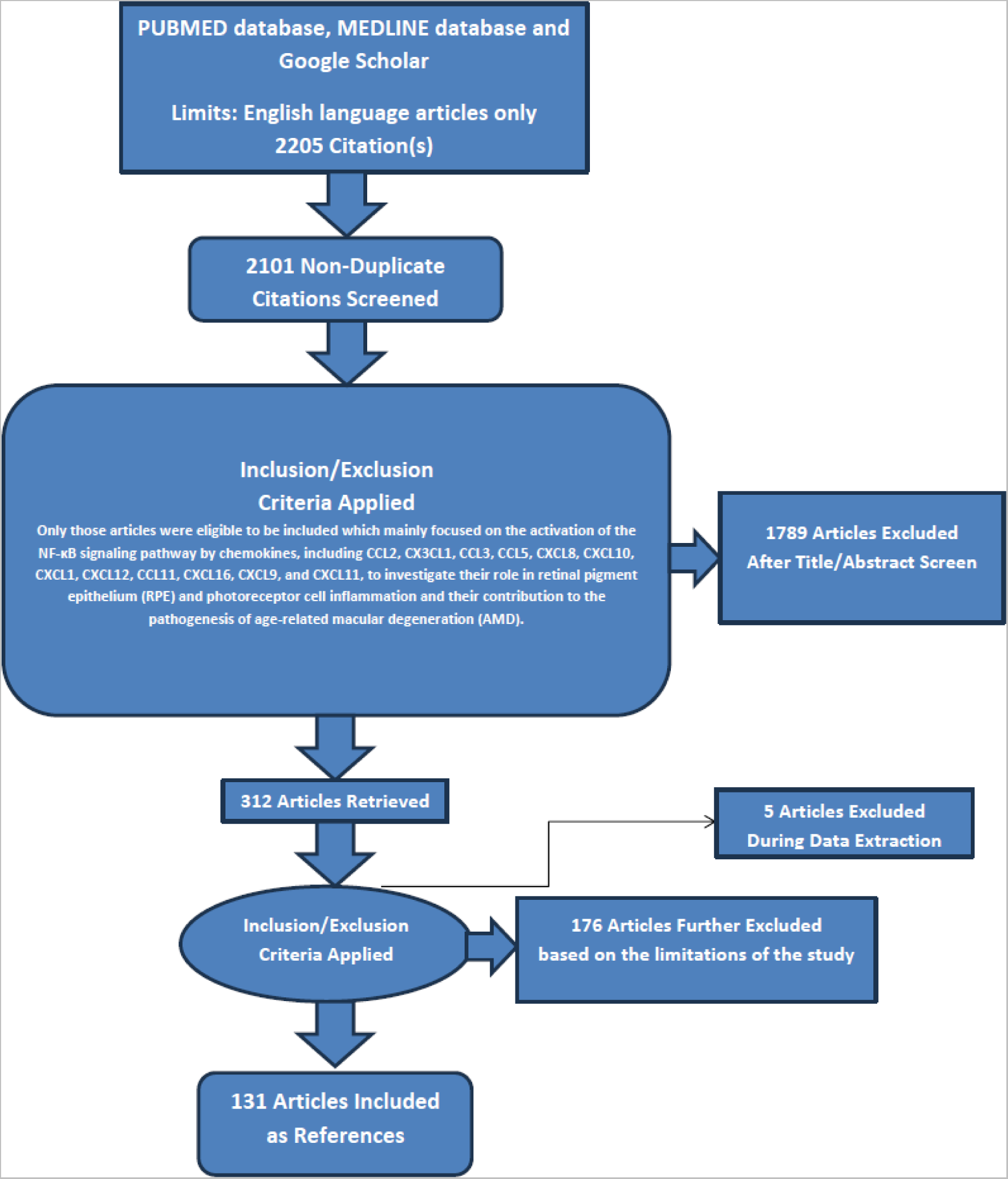
PRISMA FLOW DIAGRAM: This figure represents graphically the flow of citations in the study.

### Investigating the role of Chemokines in the Activation of the NF-κB Signaling Pathway in Retinal Pigment Epithelium (RPE) and Photoreceptor Cells

#### 1. CCL2 (MCP-1)

##### Molecular Mechanisms of CCL2 (MCP-1)-Induced NF-κB Activation

CCL2, also known as monocyte chemoattractant protein-1 (MCP-1), is a chemokine vital in recruiting monocytes, macrophages, and other immune cells to sites of inflammation. In retinal pigment epithelium (RPE) and photoreceptor cells, CCL2 contributes to the pathogenesis of age-related macular degeneration (AMD) by activating the NF-κB signaling pathway. This chemokine’s role in inflammatory processes underscores its significance in AMD development and progression [<u>8, 9</u>].

CCL2 exerts its effects by binding to the CCR2 receptor on the surface of RPE cells and photoreceptors. This binding triggers conformational changes in the receptor, leading to the activation of intracellular signaling cascades. Upon CCL2 binding, CCR2 activates associated G-proteins, which subsequently activate phospholipase C (PLC) [<u>9</u>]. PLC hydrolyzes phosphatidylinositol 4,5-bisphosphate (PIP2) into diacylglycerol (DAG) and inositol trisphosphate (IP3), where DAG activates protein kinase C (PKC) and IP3 releases calcium from intracellular stores. Additionally, the CCL2-CCR2 interaction activates the mitogen-activated protein kinase (MAPK) pathway, further contributing to NF-κB activation [<u>10, 120</u>].

The activation of NF-κB involves the phosphorylation and degradation of IκB proteins. The PKC and MAPK pathways activate IκB kinase (IKK), which phosphorylates IκB proteins, marking them for ubiquitination and degradation by the proteasome. The degradation of IκB releases NF-κB dimers, typically p65 and p50, allowing their translocation into the nucleus [<u>10, 11</u>]. In the nucleus, NF-κB binds to specific DNA sequences in the promoter regions of target genes, initiating their transcription and thereby promoting inflammatory responses associated with AMD [<u>12, 117</u>].

##### Cellular Processes Affected by CCL2 (MCP-1)-Induced NF-κB Activation

NF-κB activation leads to the upregulation of pro-inflammatory cytokines such as IL-6, TNF-α, and IL-1β, which amplify the inflammatory response and attract more immune cells to the retina. This pathway also induces the expression of additional chemokines, including CCL2 itself, creating a positive feedback loop that sustains inflammation. The persistent recruitment of immune cells and the chronic inflammation they cause contribute significantly to the pathology observed in age-related macular degeneration (AMD) [<u>13, 120</u>].

NF-κB can regulate genes involved in apoptosis, maintaining a balance between pro-apoptotic and anti-apoptotic signals. While it can induce the expression of anti-apoptotic genes like Bcl-2, chronic NF-κB activation often shifts this balance towards pro-apoptotic signals, especially in stressed or damaged retinal pigment epithelium (RPE) cells and photoreceptors. This shift towards apoptosis exacerbates cellular loss and degeneration in AMD, highlighting the dual role of NF-κB in both cell survival and cell death processes [<u>14, 120</u>].

NF-κB activation enhances the production of reactive oxygen species (ROS), contributing to oxidative stress and damage, a key factor in AMD pathogenesis. While NF-κB can also induce antioxidant genes, the chronic inflammation and elevated ROS levels often overwhelm these protective mechanisms. Additionally, NF-κB upregulates matrix metalloproteinases (MMPs), leading to the degradation of extracellular matrix (ECM) components. This degradation compromises the structural integrity of Bruch’s membrane and the RPE layer, further facilitating the progression of AMD through extracellular matrix remodeling [<u>15, 118</u>].

##### Contribution to the Pathogenesis of AMD

Sustained activation of NF-κB by CCL2 leads to chronic inflammation in the retina. This persistent inflammatory state damages retinal pigment epithelium (RPE) cells and photoreceptors, contributing to the progression of age-related macular degeneration (AMD). The ongoing inflammation perpetuates tissue damage and creates a harmful environment within the retinal layers [<u>16, 120</u>].

The inflammatory mediators and oxidative stress resulting from NF-κB activation induce apoptosis and necrosis in RPE cells and photoreceptors. This cell loss leads to the characteristic atrophy seen in dry AMD, where the degeneration of these critical cells impairs visual function. The combined effects of inflammation and oxidative stress further exacerbate cellular damage and degeneration, highlighting the destructive cycle initiated by NF-κB activation [<u>17, 120</u>].

In wet AMD, NF-κB activation promotes the expression of pro-angiogenic factors like vascular endothelial growth factor (VEGF). This facilitates the growth of abnormal blood vessels from the choroid into the retina, leading to leakage, bleeding, and further retinal damage. The upregulation of CCL2 and other chemokines by NF-κB creates a self-perpetuating cycle of inflammation and cell recruitment, exacerbating retinal damage and disease progression [<u>18, 119</u>].

#### 2. CX3CL1 (Fractalkine)

##### Molecular Mechanisms of CX3CL1 (Fractalkine)-Induced NF-κB Activation

CX3CL1, also known as fractalkine, is a unique chemokine existing in both membrane-bound and soluble forms. It plays a significant role in cell adhesion and chemotaxis, particularly in attracting microglia and monocytes. Within the retinal pigment epithelium (RPE) and photoreceptor cells, CX3CL1 has been implicated in the pathogenesis of age-related macular degeneration (AMD) through its ability to activate the NF-κB signaling pathway [<u>19, 121</u>].

CX3CL1 exerts its effects by binding to its receptor, CX3CR1, which is expressed on the surface of RPE cells, microglia, and photoreceptors. This binding initiates intracellular signaling cascades, including the activation of G-proteins, which subsequently activate phospholipase C (PLC). The PLC then hydrolyzes phosphatidylinositol 4,5-bisphosphate (PIP2) into diacylglycerol (DAG) and inositol trisphosphate (IP3). DAG activates protein kinase C (PKC), while IP3 releases calcium from intracellular stores. Additionally, the CX3CL1-CX3CR1 interaction activates the mitogen-activated protein kinase (MAPK) pathway, further contributing to NF-κB activation [<u>20, 121</u>].

The activation of NF-κB involves a series of steps, starting with the phosphorylation and degradation of IκB proteins by the IκB kinase (IKK), which is activated through PKC and MAPK pathways. The degradation of IκB releases NF-κB dimers, typically p65 and p50, from the cytoplasm, allowing their translocation into the nucleus. Once in the nucleus, NF-κB binds to specific DNA sequences in the promoter regions of target genes, initiating their transcription and contributing to the inflammatory processes associated with AMD [<u>21, 117</u>].

##### Cellular Processes Affected by CX3CL1 (Fractalkine)-Induced NF-κB Activation

NF-κB activation leads to the upregulation of pro-inflammatory cytokines such as IL-6, TNF-α, and IL-1β. These cytokines amplify the inflammatory response and attract more immune cells to the retina. Additionally, NF-κB induces the expression of additional chemokines, including CX3CL1 itself, creating a positive feedback loop that sustains inflammation [<u>22, 121</u>].

NF-κB also regulates genes involved in apoptosis, maintaining a balance between pro-apoptotic and anti-apoptotic signals. While NF-κB can induce anti-apoptotic genes such as Bcl-2, chronic activation often skews the balance towards pro-apoptotic signals, especially in stressed or damaged RPE cells and photoreceptors. This dysregulation can lead to cell death and exacerbate retinal degeneration [<u>23, 121</u>].

Activation of NF-κB enhances the production of reactive oxygen species (ROS), contributing to oxidative damage, a key factor in AMD pathogenesis. Although NF-κB can induce antioxidant genes, chronic inflammation and ROS production often overwhelm these protective mechanisms. NF-κB also upregulates matrix metalloproteinases (MMPs), which degrade components of the extracellular matrix (ECM). This degradation can compromise the structural integrity of Bruch’s membrane and the RPE layer, facilitating the progression of AMD [<u>24, 118</u>].

##### Contribution to the Pathogenesis of AMD

Sustained activation of NF-κB by CX3CL1 leads to chronic inflammation in the retina. This persistent inflammatory state damages RPE cells and photoreceptors, significantly contributing to the progression of age-related macular degeneration (AMD). The inflammatory mediators and oxidative stress resulting from NF-κB activation induce apoptosis and necrosis in RPE cells and photoreceptors, leading to the characteristic atrophy observed in dry AMD [<u>25, 121</u>].

In wet AMD, NF-κB activation promotes the expression of pro-angiogenic factors such as vascular endothelial growth factor (VEGF). This facilitates the growth of abnormal blood vessels from the choroid into the retina, resulting in leakage, bleeding, and further retinal damage. The upregulation of CX3CL1 and other chemokines by NF-κB creates a self-perpetuating cycle of inflammation and cell recruitment, exacerbating retinal damage and disease progression [<u>26, 121</u>].

The chronic inflammation driven by NF-κB leads to significant cellular damage within the retina. The continued recruitment of immune cells and the sustained production of inflammatory mediators establish a positive feedback loop, intensifying the inflammatory response. This loop exacerbates the degeneration of RPE cells and photoreceptors, highlighting the crucial role of NF-κB in the pathogenesis of both dry and wet forms of AMD [<u>27</u>,<u>119</u>].

#### 3. CCL3 (MIP-1α)

##### Molecular Mechanisms of CCL3 (MIP-1α)-Induced NF-κB Activation

CCL3, also known as macrophage inflammatory protein-1 alpha (MIP-1α), is a chemokine critical in the recruitment and activation of various immune cells, including macrophages, T cells, and dendritic cells. Within the retinal pigment epithelium (RPE) and photoreceptor cells, CCL3 significantly contributes to the pathogenesis of age-related macular degeneration (AMD) by activating the NF-κB signaling pathway. The interaction between CCL3 and its receptors CCR1 and CCR5, expressed on RPE cells and photoreceptors, plays a key role in this process [<u>28, 122</u>].

Upon binding to CCR1 and CCR5, CCL3 induces conformational changes in these receptors, initiating intracellular signaling cascades. The binding activates associated G-proteins, which subsequently stimulate phospholipase C (PLC). PLC hydrolyzes phosphatidylinositol 4,5-bisphosphate (PIP2) into diacylglycerol (DAG) and inositol trisphosphate (IP3), where DAG activates protein kinase C (PKC) and IP3 mobilizes calcium from intracellular stores. This interaction also triggers the mitogen-activated protein kinase (MAPK) pathway, further contributing to NF-κB activation [<u>29, 122</u>].

The activation of PKC and MAPK pathways leads to the phosphorylation and degradation of IκB proteins by IκB kinase (IKK). The degradation of IκB releases NF-κB dimers, typically p65 and p50, allowing their translocation into the nucleus. Once in the nucleus, NF-κB binds to specific DNA sequences in the promoter regions of target genes, initiating their transcription. This sequence of events underscores the importance of CCL3 in the inflammatory processes associated with AMD, highlighting potential therapeutic targets for intervention [<u>30</u>,<u>117</u>].

##### Cellular Processes Affected by CCL3 (MIP-1α)-Induced NF-κB Activation

NF-κB activation initiates the production of pro-inflammatory cytokines such as IL-6, TNF-α, and IL-1β, which amplify the inflammatory response and recruit additional immune cells to the retina. This activation also leads to the expression of various chemokines, including CCL3, establishing a positive feedback loop that perpetuates inflammation. The sustained inflammatory environment further exacerbates the pathological conditions in age-related macular degeneration (AMD) [<u>31, 122</u>].

In addition to its role in inflammation, NF-κB influences apoptosis by regulating genes involved in cell death. While it can promote the expression of anti-apoptotic genes such as Bcl-2, persistent NF-κB activation often shifts the balance towards pro-apoptotic signals. This shift is particularly evident in stressed or damaged retinal pigment epithelium (RPE) cells and photoreceptors, contributing to cell death and dysfunction in AMD [<u>32, 122</u>].

NF-κB activation also affects oxidative stress by enhancing the production of reactive oxygen species (ROS), which contribute to oxidative damage and are central to AMD pathology. Although NF-κB can upregulate antioxidant genes, chronic inflammation and excessive ROS production frequently overwhelm these protective responses. Additionally, NF-κB promotes the production of matrix metalloproteinases (MMPs), which degrade extracellular matrix (ECM) components. This degradation compromises the structural integrity of Bruch’s membrane and the RPE layer, facilitating AMD [<u>33, 118</u>].

##### Contribution to the Pathogenesis of AMD

Chronic activation of NF-κB by CCL3 induces a sustained inflammatory response in the retina. This persistent inflammation damages retinal pigment epithelium (RPE) cells and photoreceptors, accelerating the progression of age-related macular degeneration (AMD). The ongoing inflammatory environment contributes significantly to the degeneration observed in AMD [<u>34, 122</u>].

The inflammatory mediators and oxidative stress resulting from NF-κB activation cause apoptosis and necrosis of RPE cells and photoreceptors. This cellular damage leads to the characteristic atrophy associated with dry AMD, manifesting in a progressive loss of vision as the RPE and photoreceptor layers deteriorate [<u>35, 119</u>].

In cases of wet AMD, NF-κB activation stimulates the production of pro-angiogenic factors such as vascular endothelial growth factor (VEGF). This promotes the formation of abnormal blood vessels from the choroid into the retina, resulting in leakage, bleeding, and additional retinal damage. The continuous upregulation of CCL3 and other chemokines by NF-κB creates a positive feedback loop, perpetuating inflammation and exacerbating retinal damage and disease progression [<u>36, 122</u>].

#### 4. CCL5 (RANTES)

##### Molecular Mechanisms of CCL5 (RANTES)-Induced NF-κB Activation

CCL5, also known as RANTES (Regulated upon Activation, Normal T-cell Expressed and Secreted), is a chemokine integral to the recruitment of leukocytes during inflammation. Its role in the pathogenesis of age-related macular degeneration (AMD) is significant due to its capacity to activate the NF-κB signaling pathway within retinal pigment epithelium (RPE) and photoreceptor cells. The involvement of CCL5 in AMD underscores its potential impact on retinal inflammation and degeneration [<u>37, 123</u>].

The biological effects of CCL5 are mediated through its interaction with receptors CCR1, CCR3, and CCR5, which are present on RPE and photoreceptor cells. The binding of CCL5 to these receptors induces conformational changes that initiate intracellular signaling cascades. Specifically, CCL5-receptor interactions activate associated G-proteins, leading to the activation of phospholipase C (PLC). PLC then hydrolyzes phosphatidylinositol 4,5-bisphosphate (PIP2) into diacylglycerol (DAG) and inositol trisphosphate (IP3), resulting in the activation of protein kinase C (PKC) and the release of calcium from intracellular stores [<u>38</u>].

The signaling pathways activated by CCL5 binding subsequently influence the NF-κB signaling cascade. PKC and MAPK pathways contribute to the activation of IκB kinase (IKK), which phosphorylates IκB proteins, marking them for ubiquitination and proteasomal degradation. This degradation of IκB releases NF-κB dimers, such as p65 and p50, which then translocate to the nucleus. In the nucleus, NF-κB binds to specific DNA sequences in promoter regions, leading to the transcription of target genes associated with inflammatory responses [<u>39, 117</u>].

##### Cellular Processes Affected by CCL5 (RANTES)-Induced NF-κB Activation

NF-κB activation leads to the increased production of pro-inflammatory cytokines, including IL-6, TNF-α, and IL-1β. These cytokines enhance the inflammatory response by attracting additional immune cells to the retina. In parallel, NF-κB stimulates the expression of various chemokines, including CCL5, thereby creating a positive feedback loop that perpetuates and amplifies the inflammatory process [<u>40</u>].

NF-κB also influences apoptosis by modulating the balance between pro-apoptotic and anti-apoptotic signals. While NF-κB can activate anti-apoptotic genes such as Bcl-2, persistent activation tends to favor pro-apoptotic pathways. This shift can lead to increased cell death in stressed or damaged retinal pigment epithelium (RPE) cells and photoreceptors, exacerbating retinal pathology [<u>41, 123</u>].

Oxidative stress is another consequence of NF-κB activation, as it promotes the production of reactive oxygen species (ROS) that contribute to cellular damage. Although NF-κB can induce antioxidant genes to counteract oxidative stress, chronic inflammation and excessive ROS production frequently overwhelm these defense mechanisms. Additionally, NF-κB upregulates matrix metalloproteinases (MMPs), which degrade extracellular matrix components. This degradation compromises the structural integrity of Bruch’s membrane and the RPE layer, thereby advancing the progression of age-related macular degeneration (AMD) [<u>42, 118</u>].

##### Contribution to the Pathogenesis of AMD

Chronic activation of NF-κB, driven by CCL5, results in sustained inflammation within the retina. This persistent inflammatory environment causes significant damage to retinal pigment epithelium (RPE) cells and photoreceptors, which accelerates the progression of age-related macular degeneration (AMD) [<u>43, 119</u>].

The inflammatory mediators and oxidative stress associated with NF-κB activation contribute to both apoptosis and necrosis of RPE cells and photoreceptors. The resultant loss of these critical retinal cells manifests as the atrophy characteristic of dry AMD [<u>44</u>].

In the context of wet AMD, NF-κB activation leads to the upregulation of pro-angiogenic factors such as VEGF, promoting the growth of abnormal blood vessels from the choroid into the retina. This abnormal vascular growth results in leakage, bleeding, and further retinal damage. Additionally, the increased expression of CCL5 and other chemokines creates a self-perpetuating cycle of inflammation and cell recruitment, further exacerbating retinal damage and disease progression [<u>45, 123</u>].

#### 5. CXCL8 (IL-8)

##### Molecular Mechanisms of CXCL8 (IL-8)-Induced NF-κB Activation

CXCL8, also known as interleukin-8 (IL-8), plays a critical role in the recruitment and activation of neutrophils. Its involvement extends to retinal pigment epithelium (RPE) and photoreceptor cells, where it has been linked to the pathogenesis of age-related macular degeneration (AMD). This association is mediated through CXCL8’s capacity to activate the NF-κB signaling pathway, a key mechanism in inflammatory responses [<u>46</u>].

CXCL8 functions by binding to its receptors, CXCR1 and CXCR2, which are present on the surface of RPE cells and photoreceptors. The interaction between CXCL8 and these receptors induces conformational changes that activate intracellular signaling cascades. Specifically, CXCR1 and CXCR2 engage associated G-proteins, leading to the activation of phospholipase C (PLC). PLC then hydrolyzes phosphatidylinositol 4,5-bisphosphate (PIP2) into diacylglycerol (DAG) and inositol trisphosphate (IP3). DAG activates protein kinase C (PKC), while IP3 triggers the release of calcium from intracellular stores. Additionally, the CXCL8-CXCR interaction stimulates the mitogen-activated protein kinase (MAPK) pathway, further contributing to NF-κB activation [<u>47, 124</u>].

The activation of NF-κB is a result of several downstream events. PKC and MAPK pathways activate IκB kinase (IKK), which phosphorylates IκB proteins. This post-translational modification targets IκB for ubiquitination and degradation by the proteasome. The degradation of IκB releases NF-κB dimers, typically p65 and p50, from the cytoplasm, enabling their translocation into the nucleus. Once in the nucleus, NF-κB binds to specific DNA sequences in the promoter regions of target genes, initiating their transcription and thereby driving inflammatory responses associated with AMD [<u>48, 117</u>].

##### Cellular Processes Affected by CXCL8 (IL-8)-Induced NF-κB Activation

NF-κB activation triggers the production of pro-inflammatory cytokines, including IL-6, TNF-α, and IL-1β. These cytokines amplify the inflammatory response, drawing additional immune cells to the retina and exacerbating the inflammatory environment. Additionally, NF-κB induces the expression of chemokines, such as CXCL8, which perpetuate the inflammatory response through a positive feedback loop [<u>49, 124</u>].

NF-κB also influences the balance between pro-apoptotic and anti-apoptotic signals. It can upregulate anti-apoptotic genes, such as Bcl-2, yet chronic NF-κB activation tends to favor pro-apoptotic signals. This shift is particularly detrimental to stressed or damaged RPE cells and photoreceptors, contributing to cell death and further retinal degeneration [<u>50</u>].

The activation of NF-κB enhances the production of reactive oxygen species (ROS), leading to oxidative damage. In age-related macular degeneration (AMD), this oxidative stress is a significant pathogenic factor. Although NF-κB can induce antioxidant genes, chronic inflammation and elevated ROS levels often overwhelm these protective mechanisms. Moreover, NF-κB promotes the expression of matrix metalloproteinases (MMPs), enzymes that degrade extracellular matrix components. This degradation compromises the structural integrity of Bruch’s membrane and the RPE layer, facilitating the progression of AMD [<u>51, 118</u>].

##### Contribution to the Pathogenesis of AMD

Chronic activation of NF-κB by CXCL8 results in sustained inflammation within the retina. This ongoing inflammatory response damages retinal pigment epithelium (RPE) cells and photoreceptors, which significantly contributes to the progression of age-related macular degeneration (AMD). Persistent inflammation disrupts normal cellular function and accelerates retinal degeneration [<u>52, 124</u>].

The inflammatory mediators and oxidative stress associated with NF-κB activation induce both apoptosis and necrosis in RPE cells and photoreceptors. The resultant loss of these cells leads to the characteristic atrophy observed in dry AMD, impairing visual function and contributing to disease severity [<u>53, 119</u>].

In the context of wet AMD, NF-κB activation drives the expression of pro-angiogenic factors such as VEGF, which stimulates the growth of abnormal blood vessels from the choroid into the retina. This neovascularization leads to leakage and bleeding, exacerbating retinal damage. The upregulation of CXCL8 and other chemokines creates a positive feedback loop that perpetuates inflammation and cell recruitment, further accelerating disease progression and retinal damage [<u>54</u>].

#### 6. CXCL10 (IP-10)

##### Molecular Mechanisms of CXCL10 (IP-10)-Induced NF-κB Activation

CXCL10, also known as interferon gamma-induced protein 10 (IP-10), is a chemokine critical in immune responses by recruiting T cells, NK cells, and macrophages to sites of inflammation. In retinal pigment epithelium (RPE) and photoreceptor cells, CXCL10 has been implicated in the pathogenesis of age-related macular degeneration (AMD) through its ability to activate the NF-κB signaling pathway. This activation contributes to inflammatory processes that can exacerbate cellular damage and disease progression in AMD [<u>55, 125</u>].

CXCL10 exerts its effects by binding to its receptor, CXCR3, which is expressed on the surface of RPE cells and photoreceptors. This binding induces conformational changes in CXCR3, leading to the activation of intracellular signaling cascades. Specifically, upon CXCL10 binding, CXCR3 activates associated G-proteins, which subsequently activate phospholipase C (PLC) [<u>56</u>]. PLC then hydrolyzes phosphatidylinositol 4,5-bisphosphate (PIP2) into diacylglycerol (DAG) and inositol trisphosphate (IP3), where DAG activates protein kinase C (PKC) and IP3 releases calcium from intracellular stores. Additionally, the CXCL10-CXCR3 interaction activates the mitogen-activated protein kinase (MAPK) pathway, further contributing to NF-κB activation [<u>57</u>].

The activation of PKC and MAPK pathways leads to the activation of IκB kinase (IKK). IKK phosphorylates IκB proteins, marking them for ubiquitination and subsequent degradation by the proteasome. The degradation of IκB releases NF-κB dimers, typically p65 and p50, from the cytoplasm, allowing them to translocate into the nucleus. Once in the nucleus, NF-κB binds to specific DNA sequences in the promoter regions of target genes, initiating their transcription. This process underscores the key role of CXCL10 in mediating inflammatory responses within the RPE and photoreceptor cells, contributing to the pathophysiology of AMD [<u>58, 117</u>].

##### Cellular Processes Affected by CXCL10 (IP-10)-Induced NF-κB Activation

Inflammation plays a crucial role in the pathogenesis of AMD, primarily driven by NF-κB activation, which leads to the upregulation of pro-inflammatory cytokines such as IL-6, TNF-α, and IL-1β. These cytokines amplify the inflammatory response, attracting more immune cells to the retina, exacerbating tissue damage. NF-κB also induces the expression of additional chemokines, including CXCL10, creating a positive feedback loop that sustains chronic inflammation [<u>59, 125</u>].

NF-κB’s influence extends to the regulation of apoptosis, impacting the balance between pro-apoptotic and anti-apoptotic signals. While NF-κB can upregulate anti-apoptotic genes like Bcl-2, its chronic activation often shifts this balance towards pro-apoptotic pathways, particularly in stressed or damaged RPE cells and photoreceptors. This imbalance can lead to increased cell death, contributing to the degeneration observed in AMD [<u>60, 118</u>].

Oxidative stress is another critical factor exacerbated by NF-κB activation, which enhances the production of reactive oxygen species (ROS). Elevated ROS levels contribute to oxidative damage, a key pathogenic factor in AMD. Although NF-κB can induce antioxidant genes, the chronic inflammation and sustained ROS production often overwhelm these protective mechanisms. Additionally, NF-κB upregulates matrix metalloproteinases (MMPs), enzymes that degrade extracellular matrix (ECM) components. This degradation compromises the structural integrity of Bruch’s membrane and the RPE layer, facilitating AMD progression [<u>61, 117</u>].

##### Contribution to the Pathogenesis of AMD

Sustained activation of NF-κB by CXCL10 leads to chronic inflammation in the retina, which damages RPE cells and photoreceptors, contributing to AMD progression. This persistent inflammatory state disrupts normal cellular functions and accelerates degenerative changes within the retinal structure [<u>62</u>].

The inflammatory mediators and oxidative stress resulting from NF-κB activation induce apoptosis and necrosis in RPE cells and photoreceptors. The loss of these cells leads to the characteristic atrophy seen in dry AMD, where the retinal layers become progressively thinner and less functional due to cell death [<u>63, 125</u>].

In wet AMD, NF-κB activation promotes the expression of pro-angiogenic factors like VEGF, facilitating the growth of abnormal blood vessels from the choroid into the retina. These vessels cause leakage, bleeding, and further retinal damage. The upregulation of CXCL10 and other chemokines by NF-κB creates a self-perpetuating cycle of inflammation and cell recruitment, exacerbating retinal damage and accelerating disease progression [<u>64, 119</u>].

#### 7. CXCL1 (GRO-α)

##### Molecular Mechanisms of CXCL1 (GRO-α)-Induced NF-κB Activation

CXCL1, also known as Growth-Regulated Oncogene-alpha (GRO-α), is a chemokine crucial in inflammation and immune responses. Within the retinal pigment epithelium (RPE) and photoreceptor cells, CXCL1 has been linked to the pathogenesis of age-related macular degeneration (AMD) by activating the NF-κB signaling pathway. This activation is crucial in understanding the inflammatory processes contributing to AMD [<u>65, 126</u>].

CXCL1 exerts its biological effects by binding to the CXCR2 receptor, present on the surface of RPE cells and photoreceptors. This receptor-ligand interaction induces conformational changes in CXCR2, initiating a series of intracellular signaling cascades. One key pathway involves G-protein coupling, where the binding of CXCL1 to CXCR2 activates G-proteins that subsequently stimulate phospholipase C (PLC). PLC then hydrolyzes phosphatidylinositol 4,5-bisphosphate (PIP2) into two secondary messengers: diacylglycerol (DAG) and inositol trisphosphate (IP3). DAG activates protein kinase C (PKC), and IP3 promotes calcium release from intracellular stores, further amplifying the signal. Additionally, the CXCL1-CXCR2 interaction triggers the mitogen-activated protein kinase (MAPK) pathway, contributing to NF-κB activation [<u>66</u>].

The activation of NF-κB involves a series of molecular events, starting with the phosphorylation and degradation of IκB proteins by IκB kinase (IKK), facilitated by the PKC and MAPK pathways. This degradation frees NF-κB dimers, typically composed of p65 and p50, enabling their translocation into the nucleus. Once in the nucleus, NF-κB binds to specific DNA sequences in the promoter regions of target genes, initiating their transcription. This transcriptional activation leads to the expression of genes involved in the inflammatory response, thereby playing a significant role in the progression of AMD [<u>67, 117</u>].

##### Cellular Processes Affected by CXCL1 (GRO-α)-Induced NF-κB Activation

NF-κB activation leads to the upregulation of pro-inflammatory cytokines such as IL-6, TNF-α, and IL-1β, which amplify the inflammatory response and attract more immune cells to the retina. This heightened inflammatory state is sustained through the induction of additional chemokines, including CXCL1 itself, creating a positive feedback loop that perpetuates inflammation. The chronic inflammatory environment is a key factor in the progression of age-related macular degeneration (AMD), exacerbating retinal damage and dysfunction [<u>68, 118</u>].

NF-κB also regulates genes involved in apoptosis, influencing the balance between pro-apoptotic and anti-apoptotic signals. While NF-κB can induce the expression of anti-apoptotic genes such as Bcl-2, chronic activation often shifts the balance towards pro-apoptotic pathways, particularly in stressed or damaged RPE cells and photoreceptors. This shift can lead to increased cell death, contributing to the degeneration of retinal cells seen in AMD [<u>69</u>].

Activation of NF-κB enhances the production of reactive oxygen species (ROS), contributing to oxidative stress, a key pathogenic factor in AMD. Although NF-κB can induce the expression of antioxidant genes, the chronic inflammation and ROS production often overwhelm these protective mechanisms. Additionally, NF-κB upregulates matrix metalloproteinases (MMPs), enzymes that degrade components of the extracellular matrix (ECM). This degradation compromises the structural integrity of Bruch’s membrane and the RPE layer, further facilitating the progression of AMD [<u>70, 126</u>].

##### Contribution to the Pathogenesis of AMD

Sustained activation of NF-κB by CXCL1 leads to chronic inflammation in the retina, contributing to the progression of age-related macular degeneration (AMD). This persistent inflammatory state results in damage to retinal pigment epithelium (RPE) cells and photoreceptors, accelerating the degeneration characteristic of AMD. The continuous inflammatory response and associated oxidative stress further compromise the structural integrity of retinal cells [<u>71, 126</u>].

The inflammatory mediators and oxidative stress generated by NF-κB activation induce apoptosis and necrosis in RPE cells and photoreceptors. This cellular loss is a primary cause of the atrophy observed in dry AMD, significantly affecting retinal function and vision. The degeneration of these critical cells disrupts the normal architecture and function of the retina, exacerbating disease symptoms and advancing the progression of AMD [<u>72</u>].

In wet AMD, NF-κB activation enhances the expression of pro-angiogenic factors such as vascular endothelial growth factor (VEGF), promoting the growth of abnormal blood vessels from the choroid into the retina. These new vessels are prone to leakage and bleeding, further damaging the retina. The upregulation of CXCL1 and other chemokines by NF-κB establishes a self-perpetuating cycle of inflammation and cell recruitment, intensifying retinal damage and accelerating the progression of AMD [<u>73, 126</u>].

#### 8. CXCL12 (SDF-1)

##### Molecular Mechanisms of CXCL12 (SDF-1)-Induced NF-κB Activation

CXCL12, also known as stromal cell-derived factor-1 (SDF-1), is a chemokine involved in various physiological processes such as immune cell trafficking, angiogenesis, and tissue repair. In retinal pigment epithelium (RPE) and photoreceptor cells, CXCL12 has been implicated in the pathogenesis of age-related macular degeneration (AMD) by activating the NF-κB signaling pathway. This chemokine primarily binds to its receptor CXCR4, and to a lesser extent CXCR7, which are expressed on the surface of RPE cells and photoreceptors [<u>74, 127</u>].

Upon binding to CXCR4, CXCL12 triggers conformational changes in the receptor, leading to the activation of multiple intracellular signaling cascades. The G-protein coupling mechanism is initiated, wherein CXCR4 activates associated G-proteins that subsequently activate phospholipase C (PLC). PLC hydrolyzes phosphatidylinositol 4,5-bisphosphate (PIP2) into diacylglycerol (DAG) and inositol trisphosphate (IP3). DAG activates protein kinase C (PKC), while IP3 facilitates the release of calcium from intracellular stores. Additionally, the CXCL12-CXCR4 interaction activates the phosphoinositide 3-kinase (PI3K)/Akt pathway and the mitogen-activated protein kinase (MAPK) pathway, both of which contribute to cell survival and NF-κB activation [<u>75, 127</u>].

The activation of these signaling pathways culminates in the activation of NF-κB. The PKC, PI3K/Akt, and MAPK pathways lead to the activation of IκB kinase (IKK), which phosphorylates IκB proteins. This phosphorylation targets IκB for ubiquitination and subsequent degradation by the proteasome. The degradation of IκB releases NF-κB dimers, typically p65 and p50, from the cytoplasm, allowing them to translocate into the nucleus. In the nucleus, NF-κB binds to specific DNA sequences in the promoter regions of target genes, initiating their transcription and thereby contributing to the inflammatory processes observed in AMD [<u>76, 117</u>].

##### Cellular Processes Affected by CXCL12 (SDF-1)-Induced NF-κB Activation

Inflammation driven by NF-κB activation leads to the upregulation of pro-inflammatory cytokines such as IL-6, TNF-α, and IL-1β, amplifying the inflammatory response and attracting more immune cells to the retina.

Additionally, NF-κB induces the expression of chemokines, including CXCL12 itself, creating a positive feedback loop that sustains inflammation. This persistent inflammatory environment is a significant contributor to the pathogenesis of age-related macular degeneration (AMD) [<u>77, 127</u>].

NF-κB also plays a crucial role in regulating apoptosis by influencing the expression of genes involved in cell survival and death. While NF-κB can induce anti-apoptotic genes such as Bcl-2, chronic activation often skews the balance towards pro-apoptotic signals, particularly in stressed or damaged RPE cells and photoreceptors. This imbalance can lead to increased cell death, exacerbating the degenerative processes in AMD [<u>78, 118</u>].

Oxidative stress is another critical factor in AMD pathogenesis, with NF-κB activation enhancing the production of reactive oxygen species (ROS). The resulting oxidative damage is particularly detrimental in AMD, where oxidative stress is already a key pathogenic factor. Although NF-κB can induce antioxidant genes, the chronic inflammation and ROS production often overwhelm these protective mechanisms. Additionally, NF-κB upregulates matrix metalloproteinases (MMPs), which degrade components of the extracellular matrix (ECM). This degradation compromises the structural integrity of Bruch’s membrane and the RPE layer, facilitating AMD progression [<u>79, 127</u>].

##### Contribution to the Pathogenesis of AMD

Chronic activation of NF-κB by CXCL12 establishes a sustained inflammatory state in the retina, causing significant damage to RPE cells and photoreceptors, thereby contributing to the progression of age-related macular degeneration (AMD). This persistent inflammation disrupts cellular function and integrity, exacerbating the pathological features of AMD [<u>80, 119</u>].

The inflammatory mediators and oxidative stress induced by NF-κB activation promote apoptosis and necrosis in RPE cells and photoreceptors, leading to the characteristic atrophy observed in dry AMD. The ongoing loss of these essential cells further compromises retinal architecture and function, accelerating disease progression [<u>81</u>].

In the context of wet AMD, NF-κB activation enhances the expression of pro-angiogenic factors such as VEGF, facilitating the growth of abnormal blood vessels from the choroid into the retina. This aberrant angiogenesis results in leakage, bleeding, and additional retinal damage. Additionally, the upregulation of CXCL12 and other chemokines by NF-κB establishes a self-perpetuating cycle of inflammation and cell recruitment, further exacerbating retinal damage and advancing disease progression [<u>81, 127</u>].

#### 9. CCL11 (Eotaxin)

##### Molecular Mechanisms of CCL11 (Eotaxin)-Induced NF-κB Activation

CCL11, also known as eotaxin, is a chemokine crucial for recruiting eosinophils and other immune cells to inflammatory sites. In the context of retinal pigment epithelium (RPE) and photoreceptor cells, CCL11 plays a significant role in the pathogenesis of age-related macular degeneration (AMD). Its involvement is linked to the activation of the NF-κB signaling pathway, which is crucial in inflammatory responses [<u>82, 128</u>].

CCL11 mediates its effects by binding to the CCR3 receptor present on RPE cells and photoreceptors. This interaction induces conformational changes in CCR3, activating downstream signaling mechanisms. The binding of CCL11 to CCR3 triggers G-protein coupled receptor activation, which in turn activates phospholipase C (PLC). PLC then hydrolyzes phosphatidylinositol 4,5-bisphosphate (PIP2) into diacylglycerol (DAG) and inositol trisphosphate (IP3), with DAG activating protein kinase C (PKC) and IP3 promoting calcium release from intracellular stores. Additionally, the CCL11-CCR3 interaction stimulates the mitogen-activated protein kinase (MAPK) pathway, further contributing to NF-κB activation [<u>83, 128</u>].

The activation of NF-κB involves several key steps. PKC and MAPK pathways lead to the activation of IκB kinase (IKK), which phosphorylates IκB proteins. This phosphorylation targets IκB for ubiquitination and subsequent degradation by the proteasome. The degradation of IκB releases NF-κB dimers, such as p65 and p50, from the cytoplasm, allowing their translocation into the nucleus. Within the nucleus, NF-κB binds to specific DNA sequences in the promoter regions of target genes, initiating their transcription and contributing to the inflammatory response observed in AMD [<u>84, 117</u>].

##### Cellular Processes Affected by CCL11 (Eotaxin)-Induced NF-κB Activation

NF-κB activation leads to increased production of pro-inflammatory cytokines such as IL-6, TNF-α, and IL-1β. These cytokines amplify the inflammatory response and attract additional immune cells to the retina, exacerbating the inflammatory environment. In addition, NF-κB stimulates the expression of various chemokines, including CCL11, creating a feedback loop that perpetuates the inflammatory process and sustains the recruitment of immune cells [<u>85</u>].

NF-κB plays a critical role in regulating apoptosis by influencing the balance between pro-apoptotic and anti-apoptotic signals. While NF-κB can promote the expression of anti-apoptotic genes such as Bcl-2, chronic activation often disrupts this balance, favoring pro-apoptotic signals. This shift can lead to increased cell death among stressed or damaged RPE cells and photoreceptors, contributing to retinal degeneration [<u>86, 118</u>].

NF-κB activation also affects oxidative stress by enhancing the production of reactive oxygen species (ROS), which leads to oxidative damage and significantly contributes to age-related macular degeneration (AMD). Although NF-κB can upregulate antioxidant genes to counteract oxidative stress, persistent inflammation and ROS production often overwhelm these protective mechanisms. Furthermore, NF-κB-induced expression of matrix metalloproteinases (MMPs) contributes to extracellular matrix (ECM) remodeling, which disrupts Bruch’s membrane and the RPE layer, further facilitating the progression of AMD [<u>87, 128</u>].

##### Contribution to the Pathogenesis of AMD

Chronic activation of NF-κB by CCL11 leads to a persistent inflammatory state in the retina, resulting in ongoing damage to RPE cells and photoreceptors. This sustained inflammation accelerates the progression of age-related macular degeneration (AMD), as the inflammatory response continuously harms retinal tissues [<u>88, 128</u>].

The inflammatory mediators and oxidative stress generated through NF-κB activation drive apoptosis and necrosis of RPE cells and photoreceptors. This damage contributes to the characteristic atrophy observed in dry AMD, where the progressive loss of these cells compromises retinal function and integrity [<u>89</u>].

In wet AMD, NF-κB activation stimulates the production of angiogenic factors such as VEGF, which promotes the growth of abnormal blood vessels from the choroid into the retina. This process leads to leakage and bleeding, exacerbating retinal damage. Additionally, the upregulation of CCL11 and other chemokines by NF-κB establishes a positive feedback loop that perpetuates inflammation and cell recruitment, further aggravating retinal damage and accelerating disease progression [<u>90, 119</u>].

#### 10. CXCL16

##### Molecular Mechanisms of CXCL16-Induced NF-κB Activation

CXCL16 is a chemokine that functions both as a chemokine and a scavenger receptor, playing a significant role in immune cell recruitment and various inflammatory conditions. In retinal pigment epithelium (RPE) and photoreceptor cells, CXCL16’s activation of the NF-κB signaling pathway is crucial for understanding its involvement in age-related macular degeneration (AMD). The interaction between CXCL16 and its receptor, CXCR6, expressed on the surface of RPE cells and photoreceptors, initiates a series of intracellular signaling events [<u>91, 118</u>].

Upon binding to CXCR6, CXCL16 induces conformational changes in the receptor, which activates associated G-proteins. These G-proteins subsequently activate phospholipase C (PLC), leading to the hydrolysis of phosphatidylinositol 4,5-bisphosphate (PIP2) into diacylglycerol (DAG) and inositol trisphosphate (IP3). DAG activates protein kinase C (PKC), while IP3 triggers the release of calcium from intracellular stores. Additionally, the interaction of CXCL16 with CXCR6 activates the mitogen-activated protein kinase (MAPK) pathway, which also contributes to the activation of NF-κB [<u>92, 129</u>].

The downstream effects of this signaling cascade include the phosphorylation and degradation of IκB proteins by IκB kinase (IKK), which is activated through the PKC and MAPK pathways. This degradation process releases NF-κB dimers, such as p65 and p50, from their cytoplasmic inhibitors, allowing them to translocate into the nucleus. Once in the nucleus, NF-κB binds to specific DNA sequences in the promoter regions of target genes, leading to their transcriptional activation [<u>93, 117</u>].

##### Cellular Processes Affected by CXCL16-Induced NF-κB Activation

NF-κB activation leads to the increased production of pro-inflammatory cytokines such as IL-6, TNF-α, and IL-1β, which amplify the inflammatory response and recruit additional immune cells to the retina. This process is accompanied by the upregulation of chemokines, including CXCL16 itself, creating a positive feedback loop that perpetuates inflammation. Such chronic inflammatory signaling contributes significantly to the pathology observed in age-related macular degeneration (AMD) [<u>94, 129</u>].

In addition to inflammation, NF-κB plays a critical role in regulating apoptosis. It influences both pro-apoptotic and anti-apoptotic genes, such as Bcl-2. While NF-κB activation can induce anti-apoptotic genes, persistent activation often results in an imbalance that favors pro-apoptotic signals, leading to increased cell death in stressed or damaged retinal pigment epithelium (RPE) cells and photoreceptors [<u>95</u>].

The activation of NF-κB also impacts oxidative stress by enhancing the production of reactive oxygen species (ROS), which contributes to oxidative damage—a significant factor in AMD pathology. Despite NF-κB’s role in promoting antioxidant genes, the persistent inflammation and oxidative stress can overwhelm these protective mechanisms. Additionally, NF-κB upregulates matrix metalloproteinases (MMPs), enzymes that degrade extracellular matrix (ECM) components. This ECM remodeling disrupts Bruch’s membrane and the RPE layer, facilitating the progression of AMD [<u>96, 118</u>].

##### Contribution to the Pathogenesis of AMD

Sustained activation of NF-κB by CXCL16 leads to chronic inflammation in the retina, which damages retinal pigment epithelium (RPE) cells and photoreceptors. This persistent inflammatory state is a key factor in the progression of age-related macular degeneration (AMD), as it results in continuous cellular damage and contributes to disease advancement [<u>97</u>].

The inflammatory mediators and oxidative stress driven by NF-κB activation induce apoptosis and necrosis in RPE cells and photoreceptors. The resultant loss of these cells is a significant contributor to the atrophy observed in dry AMD, highlighting the detrimental effects of prolonged inflammatory and oxidative insults on retinal health [<u>98, 119</u>].

In cases of wet AMD, NF-κB activation also promotes the expression of angiogenic factors such as vascular endothelial growth factor (VEGF). This upregulation facilitates the growth of abnormal blood vessels from the choroid into the retina, leading to leakage, bleeding, and additional retinal damage. The positive feedback loop created by the upregulation of CXCL16 and other chemokines further exacerbates inflammation and cell recruitment, driving the progression of retinal damage and AMD [<u>99, 129</u>].

#### 11. CXCL9 (MIG)

##### Molecular Mechanisms of CXCL9 (MIG)-Induced NF-κB Activation

CXCL9, also known as monokine induced by gamma interferon (MIG), plays a crucial role in immune cell recruitment and inflammation. This chemokine is particularly significant in age-related macular degeneration (AMD) due to its involvement in the NF-κB signaling pathway within retinal pigment epithelium (RPE) and photoreceptor cells. CXCL9 exerts its effects mainly through its interaction with the CXCR3 receptor, which is expressed on RPE cells and photoreceptors, triggering downstream signaling cascades [<u>100, 130</u>].

Upon binding to CXCR3, CXCL9 activates associated G-proteins, leading to the stimulation of phospholipase C (PLC). PLC then hydrolyzes phosphatidylinositol 4,5-bisphosphate (PIP2) into diacylglycerol (DAG) and inositol trisphosphate (IP3). DAG activates protein kinase C (PKC), while IP3 causes calcium release from intracellular stores. Additionally, the CXCL9-CXCR3 interaction initiates the mitogen-activated protein kinase (MAPK) pathway, further contributing to NF-κB activation [<u>101, 130</u>].

The activation of NF-κB involves several steps. PKC and MAPK pathways activate IκB kinase (IKK), which phosphorylates IκB proteins. This phosphorylation marks IκB for ubiquitination and degradation by the proteasome. Consequently, NF-κB dimers, such as p65 and p50, are released from IκB and translocate into the nucleus. Within the nucleus, NF-κB binds to specific DNA sequences in the promoter regions of target genes, thereby initiating their transcription [<u>102, 117</u>].

##### Cellular Processes Affected by CXCL9 (MIG)-Induced NF-κB Activation

NF-κB activation triggers an increase in the production of pro-inflammatory cytokines, including IL-6, TNF-α, and IL-1β. These cytokines further amplify the inflammatory response and attract additional immune cells to the retina. Additionally, NF-κB stimulates the expression of chemokines such as CXCL9, which creates a feedback loop that perpetuates the inflammatory process [<u>103</u>].

The regulation of apoptosis by NF-κB involves both pro-apoptotic and anti-apoptotic signals. While NF-κB can promote the expression of anti-apoptotic genes like Bcl-2, chronic activation often results in a shift towards pro-apoptotic signals. This imbalance contributes to increased cell death in stressed or damaged retinal pigment epithelium (RPE) cells and photoreceptors [<u>104, 118</u>].

NF-κB activation also affects oxidative stress by enhancing the production of reactive oxygen species (ROS), leading to oxidative damage—a key factor in age-related macular degeneration (AMD). Although NF-κB can induce antioxidant genes, the persistent inflammation and oxidative stress often overwhelm these protective mechanisms. Additionally, NF-κB upregulates matrix metalloproteinases (MMPs), which degrade extracellular matrix components and disrupt structures such as Bruch’s membrane and the RPE layer, facilitating the progression of AMD [<u>105, 130</u>].

##### Contribution to the Pathogenesis of AMD

Chronic activation of NF-κB by CXCL9 leads to a persistent inflammatory state within the retina. This prolonged inflammation causes damage to retinal pigment epithelium (RPE) cells and photoreceptors, which accelerates the progression of age-related macular degeneration (AMD) [<u>130</u>].

The inflammatory mediators and oxidative stress associated with NF-κB activation result in both apoptosis and necrosis of RPE cells and photoreceptors. The consequent loss of these critical cells contributes to the atrophic changes observed in dry AMD [<u>106, 130</u>].

In cases of wet AMD, NF-κB activation facilitates the expression of angiogenic factors such as VEGF, which promotes the growth of abnormal blood vessels from the choroid into the retina. This neovascularization leads to leakage and bleeding, causing additional retinal damage. The upregulation of CXCL9 and other chemokines by NF-κB establishes a positive feedback loop that perpetuates inflammation and exacerbates retinal damage and disease progression [<u>107, 119</u>].

#### 12. CXCL11 (I-TAC)

##### Molecular Mechanisms of CXCL11 (I-TAC)-Induced NF-κB Activation

CXCL11, also known as Interferon-inducible T-cell alpha chemoattractant (I-TAC), is a chemokine involved in the recruitment of T cells and other immune cells. Its interaction with the NF-κB signaling pathway in retinal pigment epithelium (RPE) and photoreceptor cells is crucial for understanding its role in age-related macular degeneration (AMD) [<u>108, 131</u>].

CXCL11 primarily binds to its receptor CXCR3, which is expressed on RPE cells and photoreceptors. This interaction activates intracellular signaling pathways through G-proteins. The binding of CXCL11 to CXCR3 stimulates phospholipase C (PLC), leading to the hydrolysis of phosphatidylinositol 4,5-bisphosphate (PIP2) into diacylglycerol (DAG) and inositol trisphosphate (IP3). DAG activates protein kinase C (PKC), while IP3 causes the release of calcium from intracellular stores. Additionally, the CXCL11-CXCR3 interaction triggers the mitogen-activated protein kinase (MAPK) pathway, which contributes to NF-κB activation [<u>109, 131</u>].

The activation of NF-κB involves several key processes. PKC and MAPK pathways activate IκB kinase (IKK), which phosphorylates IκB proteins. This phosphorylation leads to the ubiquitination and subsequent degradation of IκB by the proteasome. The degradation of IκB releases NF-κB dimers, such as p65 and p50, allowing their translocation into the nucleus. Once in the nucleus, NF-κB binds to specific DNA sequences in the promoter regions of target genes, initiating their transcription [<u>110, 117</u>].

##### Cellular Processes Affected by CXCL11 (I-TAC)-Induced NF-κB Activation

NF-κB activation significantly increases the production of pro-inflammatory cytokines such as IL-6, TNF-α, and IL-1β. These cytokines enhance the inflammatory response and recruit additional immune cells to the retina. NF-κB also upregulates other chemokines, including CXCL11, establishing a positive feedback loop that sustains and amplifies inflammation [<u>111, 131</u>].

NF-κB plays a critical role in regulating apoptosis by influencing the balance between pro-apoptotic and anti-apoptotic signals. While NF-κB can promote anti-apoptotic genes like Bcl-2, chronic activation often shifts the balance towards pro-apoptotic signals. This shift results in increased cell death in retinal pigment epithelium (RPE) cells and photoreceptors [<u>112, 131</u>].

Increased production of reactive oxygen species (ROS) due to NF-κB activation leads to oxidative damage, a key factor in age-related macular degeneration (AMD) pathology. Although NF-κB can induce antioxidant genes, the persistent inflammation and oxidative stress often overwhelm these protective mechanisms. NF-κB also upregulates matrix metalloproteinases (MMPs), which degrade extracellular matrix components, leading to disruptions in Bruch’s membrane and the RPE layer, and thus promoting AMD progression [<u>113, 118</u>].

##### Contribution to the Pathogenesis of AMD

Sustained activation of NF-κB by CXCL11 results in persistent inflammation within the retina. This chronic inflammatory environment inflicts damage on retinal pigment epithelium (RPE) cells and photoreceptors, which contributes to the progression of age-related macular degeneration (AMD) [<u>114, 119</u>].

The inflammatory mediators and oxidative stress associated with NF-κB activation induce both apoptosis and necrosis in RPE cells and photoreceptors. The resultant loss of these critical cells is linked to the atrophic changes observed in dry AMD [<u>115, 131</u>].

In cases of wet AMD, NF-κB activation increases the expression of angiogenic factors, such as VEGF, which drives the formation of abnormal blood vessels originating from the choroid into the retina. This neovascularization causes leakage and bleeding, leading to further retinal damage. The upregulation of CXCL11 and other chemokines by NF-κB establishes a positive feedback loop that perpetuates inflammation and exacerbates retinal damage and disease progression [<u>116, 131</u>].

## Discussion

Our comprehensive investigation into the roles of various chemokines in activating the NF-κB signaling pathway in retinal pigment epithelium (RPE) and photoreceptor cells has yielded significant insights into the molecular mechanisms underpinning age-related macular degeneration (AMD). The activation of NF-κB by chemokines such as CCL2 (MCP-1), CX3CL1 (Fractalkine), CCL3 (MIP-1α), CCL5 (RANTES), CXCL8 (IL-8), CXCL10 (IP-10), CXCL1 (GRO-α), CXCL12 (SDF-1), CCL11 (Eotaxin), CXCL16, CXCL9 (MIG), and CXCL11 (I-TAC) plays a central role in driving AMD pathology through several interconnected mechanisms.

### Implications of Chemokine-Induced NF-κB Activation

Chronic inflammation plays a crucial role in the progression of age-related macular degeneration (AMD), primarily driven by the activation of NF-κB. Chemokines induce NF-κB activation, which perpetuates an inflammatory response in the retina. This sustained inflammation is marked by an increase in pro-inflammatory cytokines and chemokines, leading to progressive retinal damage. Targeting these inflammatory pathways has emerged as a promising therapeutic strategy to address the inflammatory component of AMD and potentially decelerate the disease’s advancement.

The influence of NF-κB extends to the regulation of apoptotic processes, impacting the balance between pro-apoptotic and anti-apoptotic signals. This disruption results in elevated cell death among retinal pigment epithelial (RPE) cells and photoreceptors, a hallmark of AMD, particularly in its atrophic form. Therapeutic approaches that modulate NF-κB signaling to reduce apoptosis may offer significant benefits in preserving retinal cells and sustaining visual function.

Oxidative stress, exacerbated by NF-κB-mediated upregulation of reactive oxygen species (ROS), contributes significantly to cellular damage and AMD progression. The interaction between oxidative stress and inflammation underscores the potential effectiveness of antioxidant therapies when combined with NF-κB inhibitors. Such a dual approach could address both oxidative stress and inflammation, providing a comprehensive strategy to mitigate AMD’s impact.

NF-κB activation also influences extracellular matrix (ECM) remodeling by promoting the expression of matrix metalloproteinases (MMPs), which leads to the degradation of ECM components and disruption of Bruch’s membrane. This ECM remodeling is a critical factor in AMD progression, as it impairs RPE function and fosters choroidal neovascularization (CNV). Developing interventions aimed at stabilizing ECM integrity and inhibiting MMP activity could offer therapeutic benefits for both dry and wet forms of AMD.

### Therapeutic Implications and Future Directions

Our findings reveal several promising therapeutic strategies for age-related macular degeneration (AMD). Targeting NF-κB signaling presents a viable approach to reduce chronic inflammation, potentially mitigating retinal damage and slowing disease progression. Specific inhibitors designed to block NF-κB or its upstream signaling components may provide effective treatment options for managing AMD.

Combining NF-κB inhibitors with antioxidants offers another potential strategy to address oxidative stress. This combined approach could help shield retinal cells from damage, potentially offering additional protection against AMD-related deterioration.

In addition to targeting inflammation and oxidative stress, therapies that stabilize the extracellular matrix (ECM) and inhibit matrix metalloproteinases (MMPs) hold promise. Such treatments could help prevent or reverse the retinal changes associated with AMD, thereby preserving retinal function and structure.

Future research must validate these therapeutic strategies through clinical trials and explore other potential targets within the NF-κB signaling pathway. Understanding the specific roles of various chemokines and their interactions with NF-κB will be essential for developing targeted therapies that address the complex nature of AMD. This investigation underscores the importance of chemokine-induced NF-κB activation in AMD pathogenesis and lays the groundwork for innovative approaches to managing and potentially preventing the disease.

### Key Findings

This study has provided new insights into the role of chemokine-induced NF-κB activation in the pathogenesis of age-related macular degeneration (AMD), revealing critical aspects of retinal inflammation and degeneration. Our research identified that a range of chemokines—including CCL2, CX3CL1, CCL3, CCL5, CXCL8, CXCL10, CXCL1, CXCL12, CCL11, CXCL16, CXCL9, and CXCL11—distinctly activate the NF-κB signaling pathway in retinal pigment epithelium (RPE) and photoreceptor cells. This activation initiates inflammatory responses and cellular damage, underscoring the diverse roles these chemokines play in the progression of AMD.

Chemokine-induced NF-κB activation has been shown to increase the production of pro-inflammatory cytokines and reactive oxygen species (ROS). This dual effect amplifies oxidative stress and chronic inflammation, which significantly contributes to retinal damage and the advancement of AMD. The exacerbation of oxidative stress and persistent inflammation observed in this study highlights the detrimental impact of these chemokines on retinal health.

Moreover, NF-κB activation induced by chemokines affects apoptosis in RPE and photoreceptor cells, correlating with the cell loss seen in AMD. The upregulation of matrix metalloproteinases (MMPs) due to chemokine-mediated NF-κB activation further disrupts retinal structure by promoting extracellular matrix (ECM) degradation. This degradation impairs retinal function and contributes to the progression of the disease.

Additionally, the research revealed that chemokines not only initiate NF-κB activation but also sustain a cycle of inflammation by promoting further production of chemokines. This positive feedback loop intensifies retinal damage and maintains the chronic inflammatory state characteristic of AMD. The persistence of this inflammatory cycle underscores the complexity of the disease and its underlying mechanisms.

These findings suggest that targeting chemokine-induced NF-κB activation and its downstream effects could provide novel therapeutic approaches for managing AMD. Strategies aimed at modulating inflammation, reducing oxidative stress, and mitigating ECM remodeling might offer effective ways to treat or prevent the disease. The study emphasizes the complex role of chemokines in AMD and lays the groundwork for developing targeted therapies to address its complex nature.

## Conclusion

In our investigation of the roles of various chemokines in activating the NF-κB signaling pathway within retinal pigment epithelium (RPE) and photoreceptor cells, we have uncovered several mechanisms that contribute to the development of age-related macular degeneration (AMD). Chemokines such as CCL2 (MCP-1), CX3CL1 (Fractalkine), CCL3 (MIP-1α), CCL5 (RANTES), CXCL8 (IL-8), CXCL10 (IP-10), CXCL1 (GRO-α), CXCL12 (SDF-1), CCL11 (Eotaxin), CXCL16, CXCL9 (MIG), and CXCL11 (I-TAC) each play unique yet interconnected roles in this pathological process.

Chemokines bind to their specific receptors on RPE and photoreceptor cells, initiating intracellular signaling cascades through G-protein coupled receptor activation. This engagement activates phospholipase C (PLC), which leads to the production of diacylglycerol (DAG), inositol trisphosphate (IP3), and reactive oxygen species (ROS). These intermediates contribute to the activation of NF-κB, a crucial event in the inflammatory response.

The activation of NF-κB involves the phosphorylation and subsequent degradation of IκB proteins, which allows NF-κB dimers to translocate to the nucleus. Once in the nucleus, NF-κB binds to DNA, promoting the transcription of pro-inflammatory cytokines and additional chemokines. This sustained activation of NF-κB drives a continuous inflammatory response, which is central to AMD pathogenesis.

NF-κB activation results in the upregulation of inflammatory cytokines such as IL-6, TNF-α, and IL-1β, reinforcing a chronic inflammatory environment in the retina. This persistent inflammation is a significant driver of AMD pathology. Additionally, NF-κB influences apoptosis by modulating the expression of pro- and anti-apoptotic genes, leading to increased cell death in RPE cells and photoreceptors, which contributes to retinal damage.

Furthermore, NF-κB activation enhances ROS production, leading to oxidative stress and exacerbating cellular damage. This oxidative stress plays a crucial role in the progression of AMD. Additionally, the activation of NF-κB results in the upregulation of matrix metalloproteinases (MMPs), which disrupts the extracellular matrix and impairs RPE function, further facilitating AMD progression.

In terms of AMD pathology, the chronic inflammatory state and increased apoptosis of RPE cells are characteristic of dry AMD, leading to retinal atrophy. Conversely, wet AMD is driven by sustained NF-κB activation and the expression of angiogenic factors like VEGF, which promotes choroidal neovascularization (CNV) and results in leakage and bleeding in the retina. Overall, the complex inflammatory and pathological responses orchestrated by NF-κB activation offer critical insights into the molecular mechanisms of AMD and suggest potential therapeutic targets for modulating inflammation and slowing disease progression.

## Abbreviations

AMD: Age-related Macular Degeneration
NF-κB: Nuclear Factor kappa-light-chain-enhancer of activated B cells
RPE: Retinal Pigment Epithelium
CCL2: Chemokine (C-C motif) Ligand 2 (MCP-1)
CX3CL1: Chemokine (C-X3-C motif) Ligand 1 (Fractalkine)
CCL3: Chemokine (C-C motif) Ligand 3 (MIP-1α)
CCL5: Chemokine (C-C motif) Ligand 5 (RANTES)
CXCL8: Chemokine (C-X-C motif) Ligand 8 (IL-8)
CXCL10: Chemokine (C-X-C motif) Ligand 10 (IP-10)
CXCL1: Chemokine (C-X-C motif) Ligand 1 (GRO-α)
CXCL12: Chemokine (C-X-C motif) Ligand 12 (SDF-1)
CCL11: Chemokine (C-C motif) Ligand 11 (Eotaxin)
CXCL16: Chemokine (C-X-C motif) Ligand 16
CXCL9: Chemokine (C-X-C motif) Ligand 9 (MIG)
CXCL11: Chemokine (C-X-C motif) Ligand 11 (I-TAC)
ROS: Reactive Oxygen Species
ECM: Extracellular Matrix
MMPs: Matrix Metalloproteinases

## Declarations

### Ethics declarations

Ethics approval and consent to participate

Not applicable.

### Consent for publication

Not applicable.

### Data Availability statement

All data generated or analyzed during this study are included in this article.

### Competing interests

The authors declare that they have no competing interests.

### Funding

I declare that there was not any source of funding for this research work.

## Acknowledgements

“Not applicable”.

## Authors’ Information

1. **Ifrah Siddiqui (IS)\*** is the author of the study and contributed to its conceptualization, design, and methodology, as well as the literature search and referencing. She was responsible for writing, editing, and revising the manuscript, as well as delineating the findings, results, conclusions, implications, and all other aspects of the study. IS conducted data extraction and analysis, critically evaluated every aspect of the study, ensured adherence to relevant PRISMA guidelines, and addressed study limitations and references. Additionally, she created PRISMA Flow Diagram.

She investigated the activation of the NF-κB signaling pathway by chemokines, including CCL2, CX3CL1, CCL3, CCL5, CXCL8, CXCL10, CXCL1, CXCL12, CCL11, CXCL16, CXCL9, and CXCL11, in retinal pigment epithelium (RPE) and photoreceptor cells, and their contribution to the pathogenesis of age-related macular degeneration (AMD).

Ifrah Siddiqui (IS)* holds a Bachelor’s Degree with a focus on Psychology from the University of Karachi, Pakistan. Currently, she is undertaking training/course in Health & Medicine from Harvard Medical School. She has a passion for investigating the molecular mechanisms underlying disease pathogenesis and psychological aspects of various diseases.

2. **Nabeel Ahmad Khan (NAK)** is the co-author of the study and contributed to the writing, editing and revision. He contributed to the background, results, discussion and conclusion sections of the study along with working on the findings, interpretation of the data and references.

He contributed to investigating the activation of the NF-κB signaling pathway by chemokines, including CCL2, CX3CL1, CCL3, and CCL5, in retinal pigment epithelium (RPE) and photoreceptor cells, and their contribution to the pathogenesis of age-related macular degeneration (AMD).

Nabeel Ahmad Khan (NAK) holds a Doctor of Pharmacy (Pharm.D.) degree from Dow University of Health Sciences, Karachi, Pakistan, completed in 2015, and a Master’s in Multidisciplinary Biomedical Sciences with a concentration in pharmacology from the University of Alabama at Birmingham, completed in 2022. His research interests include exploring the causal link between environmental factors and disease pathogenesis. Nabeel is dedicated to understanding how environmental influences contribute to the development and progression of various diseases, aiming to identify new therapeutic targets and strategies. Through his work, he seeks to advance the field of biomedical sciences by uncovering critical insights that can lead to improved disease prevention and treatment.

3. **Fatima Ahmad (FA)** is the co-author of the study and contributed to the writing, editing and revision. She contributed to the background, results, discussion and conclusion sections of the study along with working on the findings, interpretation of the data and references.

She contributed to investigating the activation of the NF-κB signaling pathway by chemokines, including CXCL8, CXCL10, CXCL1, and CXCL12, in retinal pigment epithelium (RPE) and photoreceptor cells, and their contribution to the pathogenesis of age-related macular degeneration (AMD).

Fatima Ahmad (FA) is currently pursuing MSc Pharmaceutical Science at Kingston University, London, UK, and holds a Doctor of Pharmacy (Pharm.D.) degree from Baqai Medical University, Karachi, Pakistan. Her ongoing research project in Kingston University is on synthesis of homoisoflavonoids for treatment of macular degeneration, and her other research interests include molecular biology and cancer chemotherapy, focusing on understanding the molecular mechanisms behind cancer progression and treatment resistance. Additionally, she explores the causal link between environmental factors and the pathogenesis of coronary artery disease. By employing advanced molecular biology techniques, Fatima aims to develop innovative therapeutic strategies for combating cancer and contributing to the advancement of precision medicine approaches that improve patient outcomes in oncology.

4. **Maham Nawaz (MN)** is the co-author of the study and contributed to the writing, editing and revision. She contributed to the background, results, discussion and conclusion sections of the study along with working on the findings, interpretation of the data and references.

She contributed to investigating the activation of the NF-κB signaling pathway by chemokines, including CCL11, CXCL16, CXCL9, and CXCL11, in retinal pigment epithelium (RPE) and photoreceptor cells, and their contribution to the pathogenesis of age-related macular degeneration (AMD).

Maham Nawaz (MN) holds a Doctor of Pharmacy (PharmD) degree from Dow University of Health and Sciences and an MPhil in Pharmacology from the University of Karachi. Her research interests are focused on stem cell diseases, exploring innovative therapeutic approaches and understanding the underlying mechanisms of disease progression.

5. **Almas Naeem (AN)** is the co-author of the study and contributed to the writing, editing and revision. She contributed to the results and conclusion sections of the study along with working on the findings, interpretation of the data and references.

She contributed to investigating the activation of the NF-κB signaling pathway by chemokines, including CCL2, CX3CL1, CCL3, and CCL5, in retinal pigment epithelium (RPE) and photoreceptor cells, and their contribution to the pathogenesis of age-related macular degeneration (AMD).

Almas Naeem (AN) holds a Doctor of Pharmacy (Pharm.D.) degree and completed an MPhil in Pharmacology from Dow University of Health Sciences. Her research interests span the fields of pharmacology, with a specific focus on in vitro and in vivo pharmacological models. She used different procedures involving cellular signal transduction mechanisms and treatment protocols as well as statistical techniques. Her previous hard work and demonstrated experience in pharmacology make her a central point in the immensely attractive field of nanotechnology.

6. **Muhammad Usaid Khalid (MUK)** is the co-author of the study and contributed to the writing, editing and revision. He contributed to the results and discussion sections of the study along with working on the findings, interpretation of the data and references.

He contributed to investigating the activation of the NF-κB signaling pathway by chemokines, including CXCL10, CXCL1, CXCL12, and CCL11, in retinal pigment epithelium (RPE) and photoreceptor cells, and their contribution to the pathogenesis of age-related macular degeneration (AMD).

Muhammad Usaid Khalid (MUK), a Pharm.D. graduate from Dow University of Health Sciences, focuses on the latest medical advancements. His valuable contributions in pharmacology, exploring different techniques and clinical trials in this area, are admirable.

7. **Usman Haroon Mirza (UHM)** is the co-author of the study and contributed to the writing, editing and revision.

He contributed to the results and conclusion sections of the study along with working on the findings, interpretation of the data and references. The author reviewed and approved the manuscript.

He contributed to investigating the activation of the NF-κB signaling pathway by chemokines, including CCL5, CXCL8, CXCL9, and CXCL11, in retinal pigment epithelium (RPE) and photoreceptor cells, and their contribution to the pathogenesis of age-related macular degeneration (AMD).

Usman Haroon Mirza holds a Doctor of Pharmacy (PharmD) degree from Dow University of Health Sciences, Karachi, Pakistan. His research interests include the molecular mechanisms of inflammation and chemokine signaling.

8. **Hamza Ali Danish (HAD)** is the co-author of the study and contributed to the writing, editing and revision. He contributed to the results and conclusion sections of the study along with working on the findings, interpretation of the data and references.

He contributed to investigating the activation of the NF-κB signaling pathway by chemokines, including CXCL1, CXCL12, CCL11, and CXCL16, in retinal pigment epithelium (RPE) and photoreceptor cells, and their contribution to the pathogenesis of age-related macular degeneration (AMD).

Hamza Ali Danish (HAD) holds a Doctor of Pharmacy (PharmD) degree from Dow University of Health Sciences. His research interests include studying the role of chemokines in inflammatory pathways and their impact on age-related macular degeneration (AMD).

9. **Syed Saqib Khalid (SSK)** is the co-author of the study and contributed to the writing, editing and revision. He contributed to the results and discussion sections of the study along with working on the findings, interpretation of the data and references.

He contributed to investigating the activation of the NF-κB signaling pathway by chemokines, including CCL3, CXCL16, CXCL9, and CXCL11, in retinal pigment epithelium (RPE) and photoreceptor cells, and their contribution to the pathogenesis of age-related macular degeneration (AMD).

Syed Saqib Khalid (SSK) is an Assistant Professor in the Department of Pharmacology at Liaquat National Hospital and Medical College, Karachi. His research interests include the pharmacological mechanisms underlying inflammatory diseases and the role of chemokines in the pathogenesis of age-related macular degeneration (AMD).

*The work and contributions of everyone have been described in detail, the order is randomized and the numbering is just for referencing purpose*.

## Notes

### Competing Interest Statement

The authors have declared no competing interest.

